# Low birthweight and prematurity, but not malaria chemoprevention, are associated with reduced pneumococcal vaccine immunogenicity in Ugandan infants

**DOI:** 10.64898/2026.05.17.26353405

**Authors:** Kenneth Musinguzi, Alyssa N Sbarra, Florian Bach, Felistas Nankya, Karen Bino Achom, Caroline Mwubaha, Patience Nayebare, Evelyn Nansubuga, Abel Kakuru, Jimmy Kizza, Zedekiah Maato, Emmanuel Arinaitwe, Kathleen Dantzler Press, Bernard Ssentalo Bagaya, Stephen Tukwasibwe, Isaac Ssewanyana, Joaniter Immaculate Nankabirwa, Moses Robert Kamya, Grant Dorsey, Saki Takahashi, Prasanna Jagannathan

## Abstract

**Background:** Malaria exposure has been hypothesized to alter immune responses to childhood vaccines, but evidence is inconsistent. We evaluated whether early-life malaria exposure and perennial malaria chemoprevention (PMC) modify antibody responses to the 10-valent pneumococcal conjugate vaccine (PCV-10) among infants in a high malaria transmission setting in eastern Uganda.

**Methods:** This study was nested within the MIC-DroP trial (NCT04978272) whereby 202 infants were selected for inclusion. Serotype-specific IgG concentrations were measured using an in-house multiplex seroassay from samples obtained at 8 and 24 weeks of age. Immunogenicity was quantified as the log_10_ fold-change in IgG concentration between the 8 and 24-week timepoints, and seroconversion as ≥0.35 μg/mL at week 24 (i.e., seropositive). Generalized estimating equation models were used to assess associations of PCV-10 immunogenicity and seroconversion with malaria exposure, malaria chemoprevention and birth outcomes.

**Results:** Among the 195 of 202 infants who completed the three-dose PCV-10 series, neither infant PMC nor malaria exposure from study enrollment to 14 weeks were associated with PCV-10 immunogenicity or seroconversion. In contrast, low birthweight (<2500g) was associated with lower immunogenicity (82% lower antibody fold-change, p=0.003) and reduced odds of seroconversion (OR=0.19, p=0.003); preterm birth (<37 weeks) showed similar associations (79% lower antibody fold-change, p=0.018; OR=0.181, p=0.009).

**Conclusion:** In this malaria-endemic setting, early-life malaria exposure and chemoprevention did not measurably alter PCV-10 antibody responses. However, low birthweight and prematurity were associated with reduced vaccine immunogenicity.

## Introduction

Pneumococcal disease, caused by the bacterium *Streptococcus pneumoniae*, is a major contributor to global child mortality^1^. Although mortality has declined substantially following introduction and scale-up of pneumococcal conjugate vaccines (PCVs)^2,3^, more recent estimates suggest that *S. pneumoniae* still accounts for approximately 300,000 deaths annually in children, primarily in sub-Saharan Africa^4^. It has been recognized there is considerable heterogeneity in vaccine immunogenicity and efficacy across geographical regions^5,6^, including for PCVs^7^. Environmental factors, nutritional status, and chronic pathogen exposure (e.g., malaria, helminths) have all been postulated to contribute to vaccine “hyporesponsiveness”^8^.

Malaria caused by the parasite *Plasmodium falciparum* is also responsible for significant morbidity and mortality. According to the World Malaria Report 2025, there were 263 million malaria cases and 597,000 deaths globally in 2023, with approximately 76% of deaths occurring in children under five years old^9^. Importantly, this age group also includes the critical window for receiving routine childhood vaccinations, under the WHO’s Expanded Programme on Immunization (EPI), including the 10-valent PCV vaccine (PCV-10)^10^.

Children in endemic areas experience malaria entomological inoculation rates of tens to hundreds per year^11,12^. Each malaria exposure results in large antigenic loads in the blood and perturbs the immunologic milieu^13^. Beyond shaping the *P. falciparum*-specific response, immune-modulating effects of *P. falciparum* infection could cause exposed infants to have difficulty mounting and sustaining effective immune responses to other antigens, including via vaccines^14^.

Malaria infection has been associated with impaired immune responses to polysaccharide vaccines^15^. While this association remains incompletely understood, chronic or repeated malaria exposure can lead to cumulative immune activation^16,17^, dysregulation of antigen-presenting cells, and perturbation of B-cell memory formation^18,19^. In addition, *in utero* exposure to malaria may also impact activation and maturation of fetal antigen-presenting cells, which could have implications for immune development in infants^20^. However, findings across studies remain inconsistent, and the additional impact of malaria chemoprevention in pregnancy (IPTp) and in infants (perennial malaria chemoprevention [PMC]^21^) on vaccine immunogenicity has not yet been well defined.

This study investigated how early-life malaria chemoprevention, malaria exposure history, and other factors affect immunogenicity and seroconversion to PCV-10 serotypes from infants enrolled in a PMC clinical trial in Busia, Uganda, a rural district with high perennial malaria transmission^22,23^. The study assessed whether early-life PMC improves PCV-10 immunogenicity and explored potential modifiers including anaemia, placental malaria, and anthropometry. Our primary hypothesis was that infants without malaria, including those receiving PMC, would have higher PCV antibody responses than those with higher malaria, or randomised to not receive PMC.

## Methods

### “Modifying Immunity in Children With DihydROartemisinin-Piperaquine” (MIC-DroP) Study

This study was nested within the MIC-DroP trial (ClinicalTrials.gov ID: NCT04978272), a randomized placebo-controlled study conducted in Busia District in eastern Uganda^24^, to evaluate the impact of early life malaria chemoprevention on malaria immunity in a high-transmission setting. Infants were born to HIV-uninfected mothers who had been randomized to three different IPTp regimens^25^. MIC-DroP enrollment took place between February 2021-June 2023 during which 924 infants between 4 and 8 weeks of age were randomized to receive PMC with monthly dihydroartemisinin-piperaquine (DP) from 8 weeks to either 52 or 104 weeks (n=616) or placebo (n=308). Malaria exposure was assessed through active and passive surveillance. From enrollment, finger-prick blood samples were collected every 4 weeks for detection of parasitemia by microscopy and quantitative PCR (qPCR). Children were also evaluated at the study clinic for all acute illnesses outside of scheduled visits, with blood samples obtained for microscopy and qPCR whenever fever was present. Each thick smear was independently read by two experienced microscopists, with discrepancies resolved by a third reader. Malaria exposure at any time from enrollment to 14-weeks-old (hereafter peri-vaccination window) was defined in two ways: clinical malaria as presence of parasites on blood smear with concurrent fever (axillary temperature ≥38.0°C), and asymptomatic parasitemia as detection of parasitemia by microscopy or qPCR in absence of fever.

### Study design

A total of 202 children were randomly selected from all MIC-DroP participants to ensure representation from randomization arms. Because primary immunogenicity outcomes were measured at 24 weeks (i.e., when both PMC arms received identical monthly chemoprevention), infants were combined into a single ‘PMC exposed’ group for analysis. Plasma samples collected at 8 and 24 weeks of age were selected for testing in this study (n=404 samples total).

PCV-10 is administered through Uganda’s EPI program at 6, 10, and 14 weeks of age. In this cohort, the median age at receipt of the third PCV-10 dose was 16.1 weeks (IQR 15.0–17.9), with a mean age of 18.8 weeks, indicating that a subset of children received third doses later than scheduled. The PCV formulation used in Uganda contains serotypes 1, 4, 5, 6B, 7F, 9V, 14, 18C, 19F, and 23F^3^. Vaccination status was determined from vaccination cards and parental recall recorded at study visits. Inclusion in the primary analysis required documented receipt of all three PCV-10 doses; n=195 of 202 infants met this criterion.

Sensitivity analyses assessed robustness by restricting analyses to only children with all three PCV-10 doses documented on their vaccination card (n=156) and by expanding analyses to all children who received at least one PCV-10 dose from either vaccination card or parental recall (n=202).

### Multiplex bead assay

To measure PCV-10 antibody responses, IgG concentrations specific to each pneumococcal serotype listed above were quantified using an in-house multiplex bead-based immunoassay (Magpix; Luminex Corporation, USA). Luminex beads were conjugated to *S. pneumoniae* polysaccharides as described previously^26^. Briefly, polysaccharides (1 mg/mL, American Type Culture Collection (USA)) were activated with 4-(4,6-dimethoxy [1,3,5] triazin-2-yl)-4-methyl-morpholinium (DMTMM) for 40-60 minutes at room temperature^27^, purified using Sephadex G-25 columns to remove excess reagent, and incubated overnight with microspheres in the dark. Conjugated beads were washed with PBST and stored in blocking buffer at 4°C.

Plasma samples were diluted 1:500 in assay buffer (PBS containing 10% bovine serum albumin, 0.05% sodium azide, and cell wall polysaccharide [Statens Serum Institut, Denmark]) to minimize non-specific binding and incubated with 375 serotype-specific antigen-coupled beads per well. After incubation with diluted plasma, beads were washed and phycoerythrin-conjugated goat anti-human IgG (Jackson ImmunoResearch, USA) was added. Following a final wash, median fluorescence intensities (MFIs) were obtained on a Magpix instrument. MFI values were converted to IgG concentrations (µg/mL) using a four-parameter logistic model fit to 11 serial dilutions of the internationally-recognized pneumococcal reference serum (007SP)^28^.

### Ascertainment of exposures

In addition to febrile malaria, asymptomatic parasitemia, and PMC treatment allocation other exposures assessed included maternal gravidity, IPTp arm, placental malaria, sex, birthweight, gestational age, sickle cell status (via HBS genotype), and anemia. Maternal gravidity was classified as primigravida or multigravida. IPTp arm was defined by a randomized regimen from the MIC-DroP parent study (sulfadoxine pyrimethamine [SP], dihydroartemisinin-piperaquine [DP], or SP+DP)^25^. Infant PMC exposure was defined according to MIC-DroP randomization arm (DP or placebo)^24^. Low birth weight (LBW) was defined as <2500 grams, and preterm birth was defined as having a gestational age less than 37 weeks at time of birth. Placental malaria was determined histologically using standard criteria and classified as either active (parasites present) or past infection (malaria pigment without parasites)^29^. HBS genotype was classified as HbAA (normal hemoglobin) or HbAS (sickle cell trait)^30^. Anemia was defined as haemoglobin <10 g/dL^31^ at 8 weeks.

### Statistical analysis

Using preliminary data, the mean log□□ antibody concentration for PCV serotype 5 at week 24 was 2.99 with a SD of 0.115. Assuming 100 participants per group, a two-sided α=0.05, and 80% power, the study was powered to detect a minimum difference of approximately 0.045 log□□ antibody concentration units between DP versus placebo groups at week 24. The primary outcome was PCV-10 immunogenicity, defined as the log□□ fold-change in serotype-specific IgG concentrations between 8 and 24 weeks. The secondary outcome was seroconversion, defined as achieving the WHO-recommended IgG threshold of ≥0.35 µg/mL^32^ at 24 weeks. To account for correlation of multiple serotype responses within a child, generalized estimating equation (GEE) models with individual-level clustering were used to estimate differences in immunogenicity and sero-conversion. Serotype-specific IgG responses were treated as repeated measures nested within individuals, enabling estimation of population-level effects across serotypes. Statistically-significant pooled associations were followed by serotype-specific GEE analyses to assess heterogeneity. All analyses were performed using R version 4.4.1. GEE models were fitted using the *geepack* package, specifying an exchangeable working correlation structure to account for within-individual correlation^33^. Computational code can be found at https://github.com/musinguzi-kenneth/PCV-10-analysis.

### Ethics

Ethical approval was obtained from institutional review boards of Stanford University, Makerere University School of Biomedical Sciences Research and Ethics Committee, and the Uganda National Council for Science and Technology. Written informed consent was obtained from parents or legal guardians of all participants before enrollment. The study was conducted in accordance with established ethical guidelines for human subjects’ research.

## Results

### Demographics and clinical characteristics

195 infants had documented completion of three-dose of PCV-10 (**Table 1**). 5.1% were classified as LBW (<2500 g) and 4.1% as preterm (<37 weeks). Symptomatic malaria during the peri-vaccination window was rare at 1%, while asymptomatic parasitemia detected by qPCR in the peri-vaccination window occurred in 18% of infants. Among children with at least 1 PCV-10 dose noted on vaccination cards (n=174), 85.6% (n=149) had received at least 1 PCV-10 dose by their 8 week visit. The number of PCV-10 doses received is summarized in **Table S1**. The median age of receipt of the third dose of PCV-10 among children with 3 doses documented on vaccine cards was 16.1 weeks.

**Table 1.** Demographic and clinical characteristics of infants (n = 195) included in the immunogenicity substudy nested within the MIC-DroP trial in Busia District, Uganda.

| Variable | Strata | Total (n = 195) |
| --- | --- | --- |
| Sex (n, %) | Male | 92 (47.0%) |
|  | Female | 103 (53.0%) |
| Birth weight (n, %) | Low (<2500 g) | 10 (5.1%) |
| | Not low birthweight (≥2500 g) | 185 (94.9%) |
| Anemia status at 8 weeks (n, %) | Yes | 133 (68.0%) |
|  | No | 62 (32.0%) |
| Clinical malaria (BS positive & symptomatic) between enrollment and 14 weeks (n, %) | Yes | 2 (1.0%) |
|  | No | 193 (99.0%) |
| Parasitemia (qPCR > 0.001 parasites/μL) between enrollment and 14 weeks (n, %) | Yes | 36 (18.0%) |
|  | No | 159 (82.0%) |
| Parasitemia between enrollment and 6 weeks | Yes | 6 (3.0%) |
|  | No | 153 (78.5%) |
|  | Not enrolled prior to 6 weeks | 36 (18.5%) |
| Gestational age (n, %) | Preterm (<37 weeks) | 8 (4.0%) |
| | Not preterm (≥37 weeks) | 187 (96.0%) |
| Gravidity of mother (n, %) | Primigravida | 42 (22.0%) |
|  | Multigravida | 153 (78.0%) |
| HbS genotype (sickle cell) (n, %) | HbAS | 40 (21.0%) |
|  | HbAA | 154 (79.0%) |
| Maternal chemoprevention arm (n, %) | SP | 71 (36.0%) |
|  | DP | 66 (34.0%) |
|  | SP+DP | 58 (30.0%) |
| PMC regimen (n, %) | No DP | 102 (52.3%) |
|  | DP | 93 (47.7%) |
| Placental malaria (PM) (n, %) | PM- | 109 (59.0%) |
|  | PM+ | 76 (41.0%) |

### PCV-10 antibody responses increase from weeks 8 to 24

Serotype-specific pneumococcal IgG concentrations were measured at 8 and 24 weeks of age (**Figure 1**). For all vaccine serotypes, median IgG concentrations were higher at 24 weeks than at 8 weeks. At 8 weeks, antibody concentrations were generally low with wide interquartile ranges. Seroprevalence at week 8 ranged from 8.2% for serotype 6B to 65.1% for serotype 14, and seroprevalence at week 24 ranged from 76.9% for serotype 23F to 92.8% for serotype 7F. The magnitude of increase (i.e., immunogenicity) varied between serotypes. Across serotypes, immunogenicity ranged from 1.83 log□□ fold-change (serotype 14) to 4.33 log□□ fold-change (serotype 7F).

**Figure 1:**
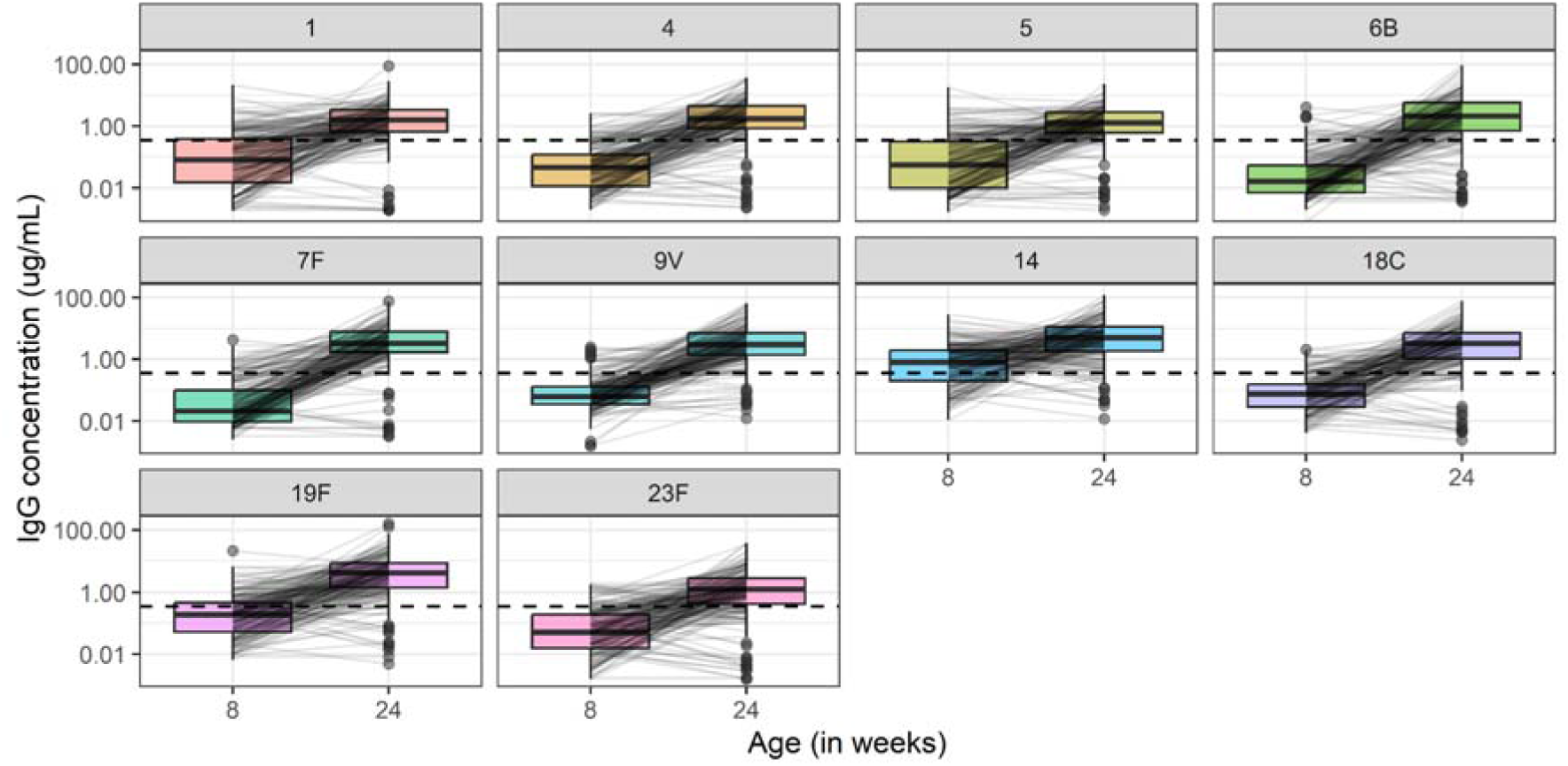
Serotype-specific pneumococcal IgG concentrations at 8 and 24 weeks of age following PCV-10 vaccination. Box plots show the distribution of IgG concentrations (µg/mL) for each PCV-10 serotype at each time point. The horizontal line within each box represents the median. The horizontal dashed line on each box represents the WHO recommended IgG seroprotective cut-off of ≥0.35 µg/mL. IgG concentrations were measured using a Luminex-based multiplex immunoassay, and these are shown on a log□□ scale. Trends illustrate variability in antibody acquisition and persistence across serotypes.

### Low birthweight and preterm birth associated with lower immunogenicity and seroconversion

There were no significant pooled associations with immunogenicity or seroconversion observed for infant PMC (DP vs. placebo) or presence of parasitemia during the peri-vaccination period (**Table 2**, serotype-specific results in **Table S2** and **Table S3**). Parasitemia during the peri-vaccination period was neither associated with higher odds of seroconversion (OR=2.31, p=0.077), nor immunogenicity (2% lower fold-change, p=0.983). In addition, no significant pooled associations with immunogenicity or seroconversion were observed for infant sex, sickle cell trait, maternal gravidity, placental malaria, anaemia, and maternal IPTp regimen (**Table 2**). In the parent cohort to MIC-DroP, IPTp regimens of DP or DP+SP were both associated with significantly reduced burden of malaria in pregnancy, and reduced burden of placental malaria^25^.

**Table 2:**
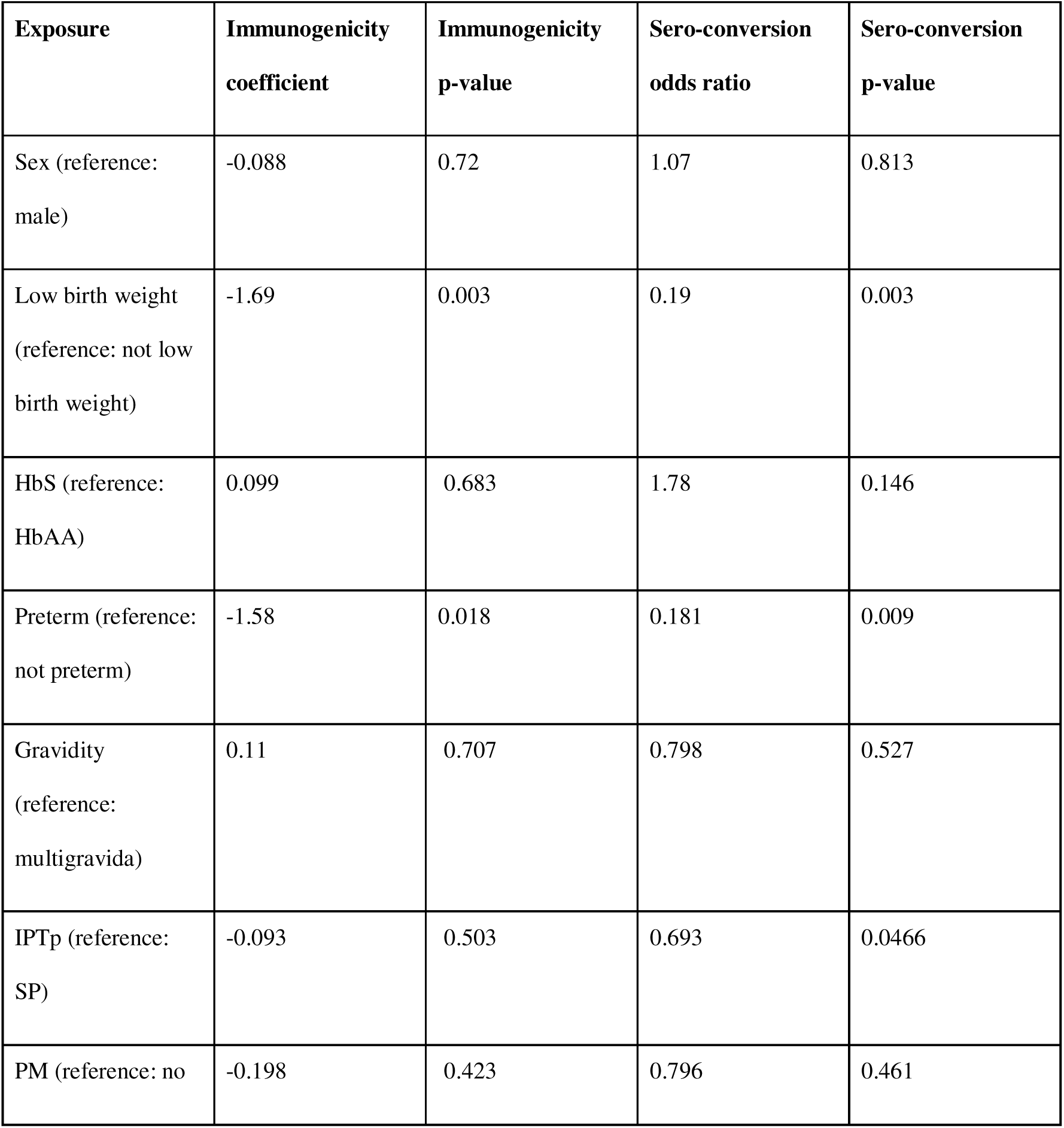
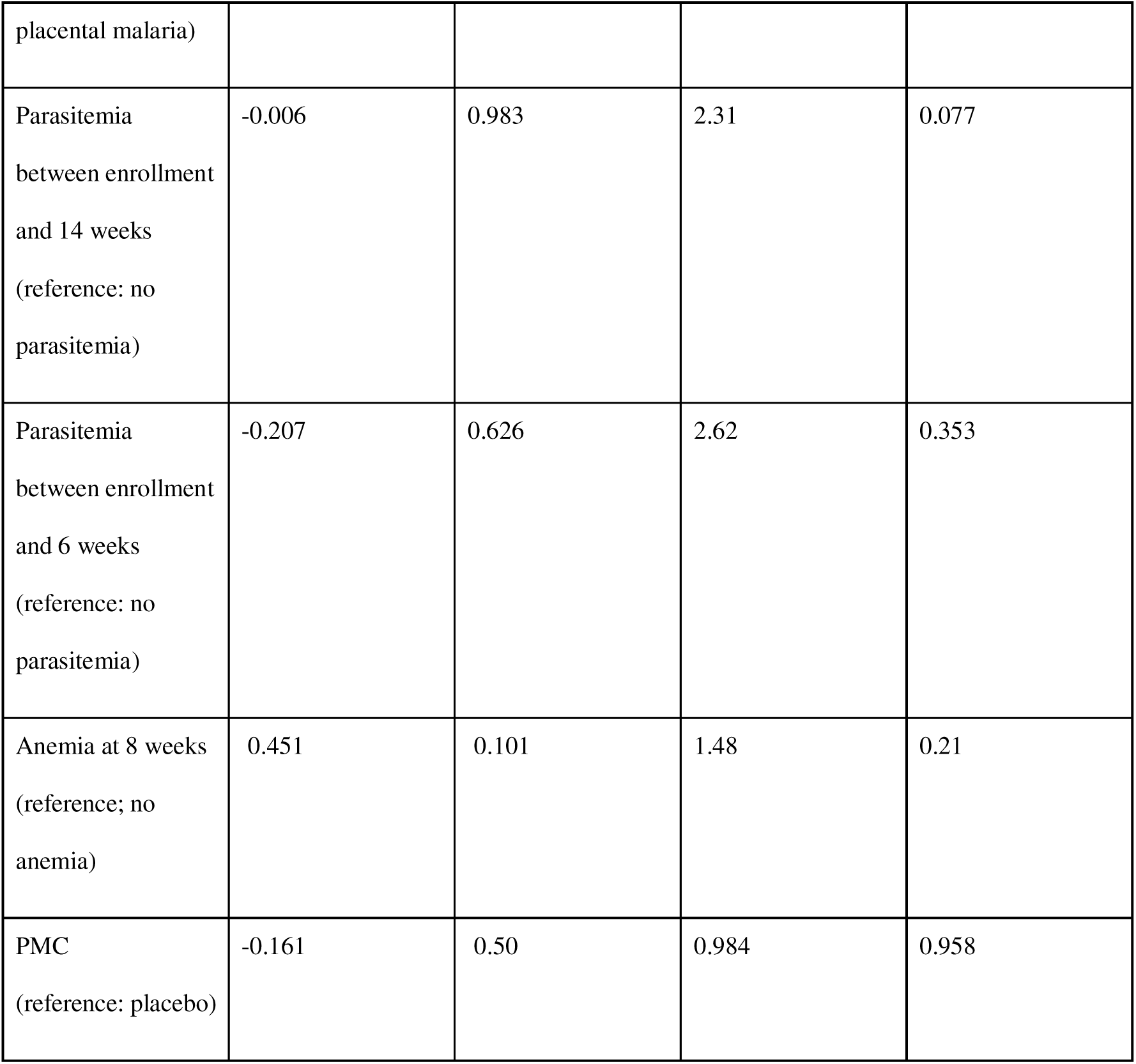
Pooled generalized estimating equation (GEE) results for PCV-10 immunogenicity and seroconversion. Immunogenicity was defined as the log□□ fold-change in serotype-specific pneumococcal IgG concentrations between 8 and 24 weeks of age, and seroconversion was defined as achieving a serotype-specific IgG concentration ≥0.35 µg/mL at 24 weeks of age. Pooled GEE models treated serotype-specific responses as repeated measures within participants and were clustered at the individual level. Coefficients represent mean differences in log□□ fold-change IgG concentrations, and odds ratios represent the odds of seroconversion. Statistical significance was assessed at α = 0.05 (HbS = sickle hemoglobin, IPTp = intermittent preventive treatment in pregnancy, PM = placental malaria, PMC = perennial malaria chemoprevention).

In contrast, low birthweight and preterm birth were each associated with poorer PCV-10 responses. Low birthweight (i.e., <2500 g at birth) was associated with lower immunogenicity (82% lower fold-change, p=0.003) and reduced odds of seroconversion (OR=0.19, p=0.003), compared with infants with birth weights of 2500 g or greater. Similarly, infants born preterm (i.e., <37 weeks gestational age) had lower immunogenicity (79% lower fold-change, p=0.018) and reduced odds of seroconversion (OR=0.181, p=0.009), compared with infants not born at or after 37 weeks of gestational age (**Table 2**).

For both low birthweight and preterm birth, additional serotype-specific GEE analyses were undertaken to assess heterogeneity across individual PCV-10 serotypes. For immunogenicity (**Table 3**), low birthweight was associated with significantly lower log□□ fold-change IgG concentrations for 6 out of 10 serotypes after adjustment for multiple comparisons. Preterm birth was associated with reduced immunogenicity for 4 out of 10 serotypes. Low birthweight was associated with significantly lower odds of seroconversion for 7 out of 10 serotypes, while preterm birth was associated with reduced odds of seroconversion for 6 out 10 serotypes (**Table 4**, **Figure 2**, **Figure S1**).

**Figure 2:**
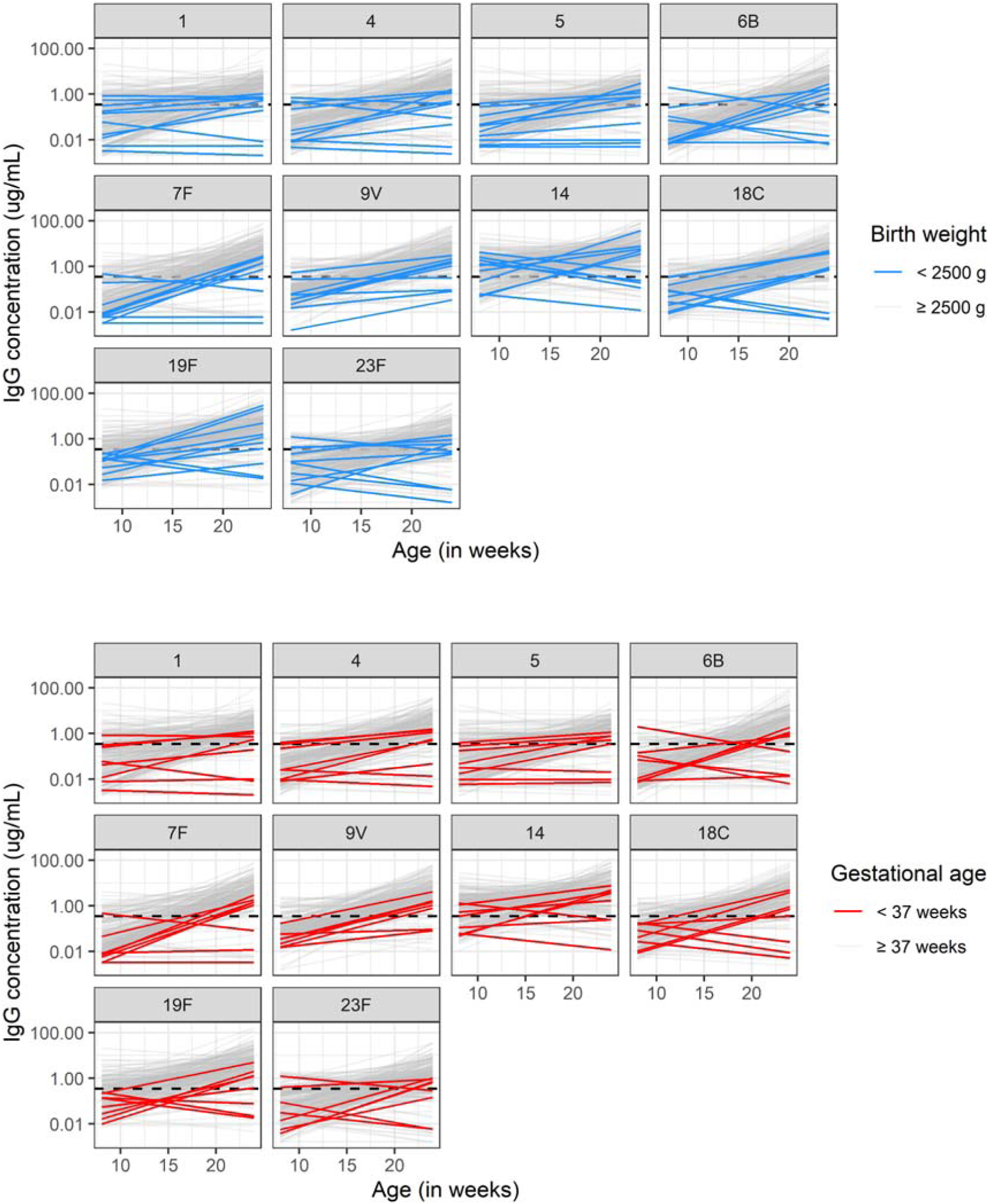
Longitudinal serotype-specific pneumococcal IgG responses following PCV-10 vaccination, stratified by birthweight (top) and gestational age (bottom). Lines represent individual IgG concentrations for each PCV-10 serotype measured at 8 and 24 weeks of age. Panels highlight differences by <2500 g (low birthweight) versus birthweight ≥2500 g and by <37 weeks (preterm) versus term birth ≥37 weeks. The horizontal dashed line on each box represents the WHO recommended IgG seroprotective cut-off of ≥0.35 µg/mL.

**Table 3:** Immunogenicity expressed as log׈׈ fold-change in serotype-specific pneumococcal IgG concentrations between 8 and 24 weeks of age following PCV-10 vaccination, comparing low birthweight infants to preterm birth infants. Coefficients are from pooled generalized estimating equation (GEE) models clustered by child, with serotype-specific estimates shown. Negative coefficients indicate lower fold-increase in IgG concentrations among low birthweight infants and preterm birth infants. P-values are adjusted for multiple comparisons across serotypes using the Benjamini-Hochberg (BH) method.

| Serotype | Low birthweight<br>coefficient | Low<br>birthweight | Preterm birth<br>coefficient | Preterm birth<br>adjusted p- |
| --- | --- | --- | --- | --- |

|  |  | adjusted p-value |  | value |
| --- | --- | --- | --- | --- |
| 1 | -2.17 | <0.001 | -2.13 | 0.001 |
| 4 | -2.39 | 0.002 | -2.09 | 0.001 |
| 5 | -1.26 | 0.028 | -1.78 | 0.001 |
| 6B | -2.33 | 0.041 | -2.99 | 0.023 |
| 7F | -1.26 | 0.222 | -1.29 | 0.271 |
| 9V | -1.12 | 0.031 | -0.626 | 0.269 |
| 14 | -1.15 | 0.267 | -0.367 | 0.646 |
| 18C | -1.73 | 0.068 | -1.91 | 0.131 |
| 19F | -0.896 | 0.267 | -1.32 | 0.269 |
| 23F | -2.54 | 0.003 | -1.26 | 0.278 |

**Table 4:**
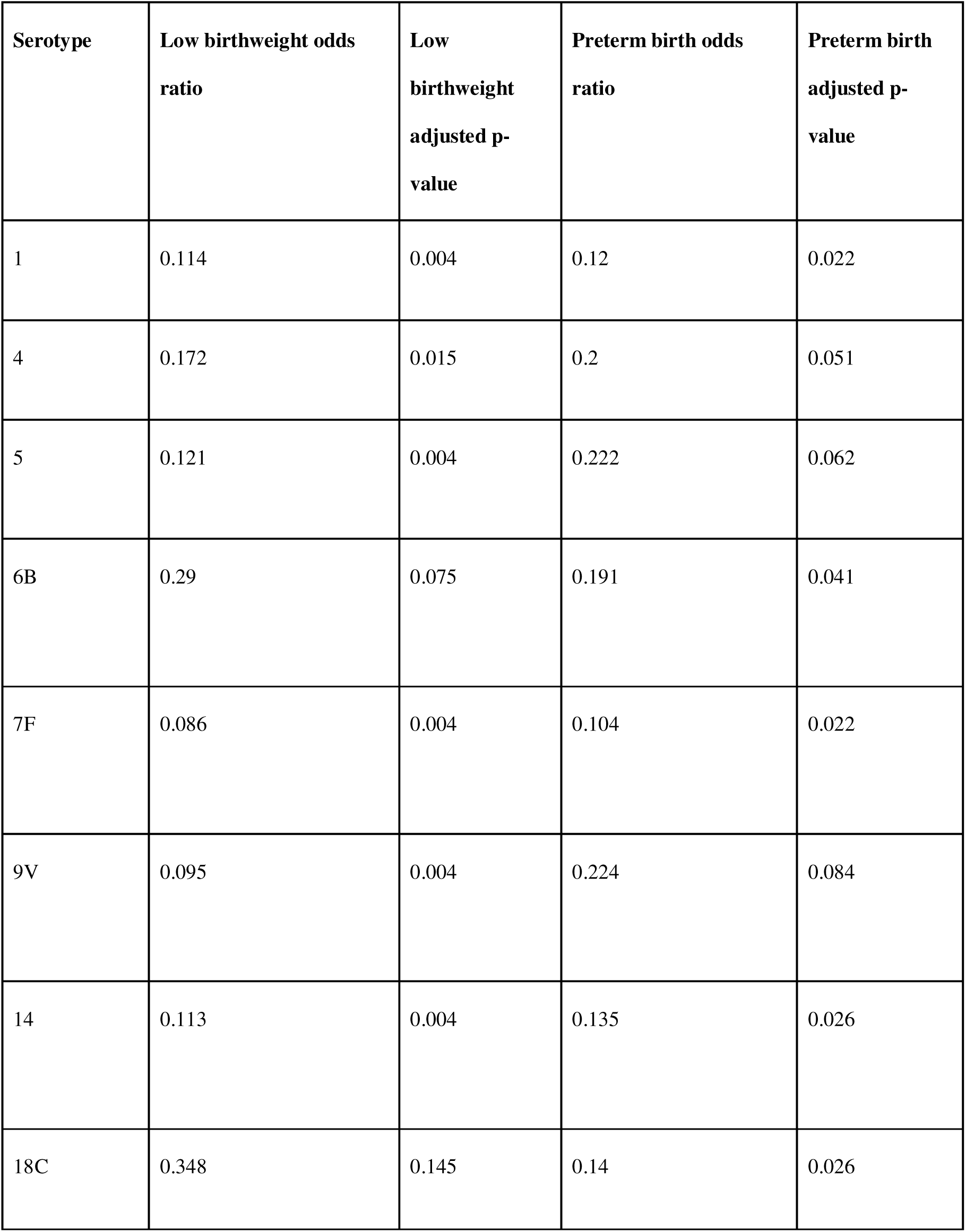
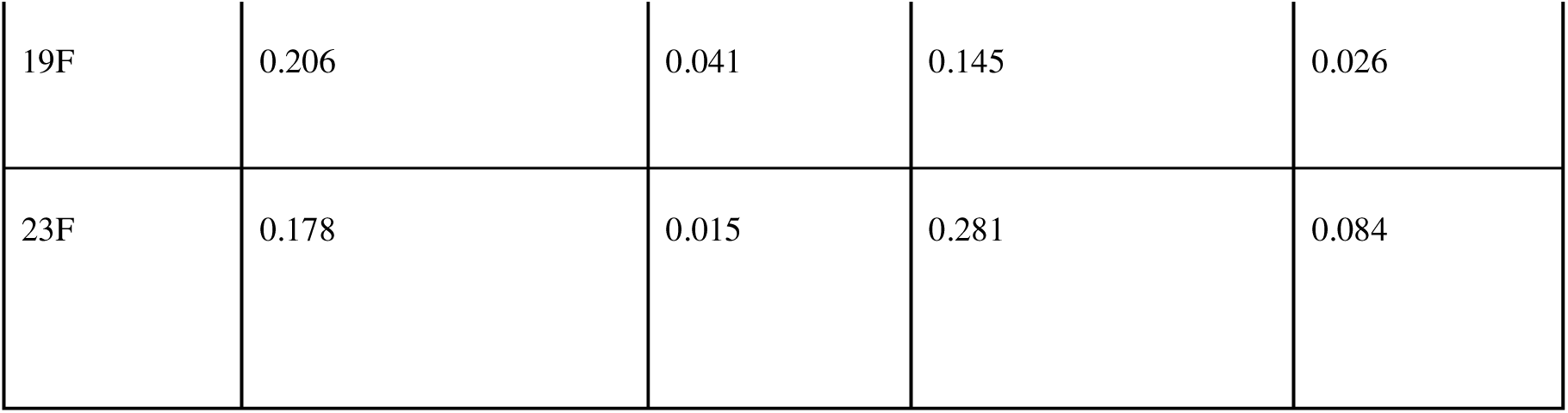
Serotype-specific seroconversion following PCV-10 vaccination according to early-life exposures. Odds ratios for seroconversion (defined as pneumococcal IgG concentration ≥ 0.35 µg/mL at 24 weeks of age) were estimated using pooled generalized estimating equation (GEE) logistic regression models clustered by child. Results are shown for low birthweight and preterm birth at the time of vaccination, with serotype-specific estimates presented. Odds ratios < 1 indicate reduced odds of seroconversion among exposed infants compared with the unexposed reference group. P-values were adjusted for multiple comparisons across serotypes using the Benjamini-Hochberg (BH) method.

| Serotype | Low birthweight odds ratio | Low birthweight adjusted p-value | Preterm birth odds ratio | Preterm birth adjusted p-value |
| --- | --- | --- | --- | --- |
| 1 | 0.114 | 0.004 | 0.12 | 0.022 |
| 4 | 0.172 | 0.015 | 0.2 | 0.051 |
| 5 | 0.121 | 0.004 | 0.222 | 0.062 |
| 6B | 0.29 | 0.075 | 0.191 | 0.041 |
| 7F | 0.086 | 0.004 | 0.104 | 0.022 |
| 9V | 0.095 | 0.004 | 0.224 | 0.084 |
| 14 | 0.113 | 0.004 | 0.135 | 0.026 |
| 18C | 0.348 | 0.145 | 0.14 | 0.026 |
| 19F | 0.206 | 0.041 | 0.145 | 0.026 |
| 23F | 0.178 | 0.015 | 0.281 | 0.084 |

Because IPTp regimen, parasitemia, and infant PMC directly reflect malaria exposure during the period of immune priming, we further explored serotype-specific models for these exposures to assess potential heterogeneity in antibody responses across individual PCV-10 serotypes, among infants that received all 3 doses (n=195). After accounting for multiple comparisons, no statistically significant patterns emerged across individual serotypes (**Tables S2–S3**).

### Sensitivity analyses

We conducted several additional analyses to test the robustness of our main findings. First, we restricted the analysis to infants with three PCV doses documented on vaccination cards (i.e., instead of vaccination history obtained from both vaccination cards and parental recall, n=156) to reduce the possibility of misclassification arising from parental recall (**Table S4**). Low birthweight continued to show a strong association with reduced PCV-10 responses, including lower immunogenicity and reduced odds of seroconversion; preterm birth was also associated with lower odds of seroconversion. Similarly, malaria exposure and infant PMC were not associated with either immunogenicity or seroconversion.

Second, we repeated pooled GEE analyses among all children who had received at least one dose of PCV-10 (i.e., with history obtained from either vaccination card or parental recall n=202; **Table S5**). The findings were again consistent: low birthweight and preterm birth both remained the only exposures associated with lower immunogenicity and reduced odds of seroconversion.

Then, among children with the date of PCV-10 receipt documented on vaccination cards, we explored use of time since vaccination as an additional covariate in all univariate GEE models of immunogenicity and seroconversion (**Table S6, Figure S2**). Inclusion of time since vaccination did not alter the overall pattern of associations observed in primary analyses. Low birthweight remained strongly associated with reduced immunogenicity and lower odds of seroconversion, and preterm birth showed reduced odds of seroconversion. Anaemia at 8 weeks was associated with higher immunogenicity in these models, although no corresponding association with seroconversion was observed.

Finally, also among children with the date of PCV1 receipt documented on vaccination cards, we examined univariate GEE analyses of parasitemia with different peri-vaccination windows defined based on individual-level vaccination timing, including parasitemia between enrollment and receipt of the first PCV dose, between enrollment and receipt of the third dose, and between first and third PCV doses (**Table S7**). Across all definitions, parasitemia was not significantly associated with either immunogenicity or seroconversion. These findings were consistent with primary analyses using fixed age-based exposure windows.

## Discussion

We investigated whether malaria exposure during early infancy and PMC modified antibody responses to PCV-10 in a high-transmission setting. Associations between peri-vaccination parasitemia, infant PMC, maternal IPTp, placental malaria, and host characteristics were examined in relation to immunogenicity and seroconversion between 8 and 24 weeks of age. In this study, we did not find evidence that early-life malaria exposure or infant perennial PMC altered immunogenicity or seroconversion to PCV-10 antigens. Instead, differences in vaccine responses were seen among infants born with low birthweight or prematurely. These infants had lower immunogenicity and were less likely to seroconvert by 24 weeks of age.

Parasitemia detected by qPCR during the peri-vaccination period was not associated with reduced immunogenicity or lower odds of seroconversion in pooled analyses. Similarly, infants randomized to monthly DP did not show stronger antibody responses than those receiving placebo. Although there was a non-significant trend toward higher odds of seroconversion among infants with parasitemia, this pattern was not supported by differences in antibody fold-change. Our findings differ from reports in settings where malaria exposure occurred at the time of vaccination. A systematic review and meta-analysis concluded that parasite infections present at the time of vaccination are associated with lower vaccine responses^8^. Prior work in Uganda has suggested that maternal and infant infections can modulate early vaccine responses in an antigen-specific and context-dependent manner, though these immune suppression effects are not uniform across different vaccine antigens (e.g., BCG versus tetanus toxoid).^34^ However, other controlled trials showed that reducing malaria exposure through chemoprophylaxis did not change vaccine responses to measles, diphtheria, tetanus and pertussis^35^. The timing of chemoprevention relative to vaccination may have limited its capacity to influence vaccine responses. Infants received their first dose of DP at 8 weeks of age, whereas the first dose of PCV-10 is administered at 6 weeks. Thus, the initial priming dose of PCV-10 preceded the initiation of chemoprevention. If malaria exposure influences early priming events, the delayed start of PMC may have limited its capacity to modify subsequent antibody acquisition. This temporal sequence may partly explain the absence of a measurable PMC effect on PCV-10 responses in our cohort.

Further, maternal IPTp regimen and placental malaria were also not associated with differences in infant immunogenicity or seroconversion. Because IPTp regimens including monthly DP reduce malaria in pregnancy and placental malaria^29^, thereby limiting fetal exposure to malaria antigens and potential *in utero* immune modulation, we hypothesized that improved control of maternal malaria might enhance early-life vaccine responses. However, in this cohort, modification of *in utero* malaria exposure did not translate into measurable differences in PCV-10 immunogenicity. Studies have shown that different forms of prenatal malaria exposure are associated with heterogeneous alterations in fetal immune responses, with substantial inter-individual variability^36^. In addition, in utero exposure may induce either immune activation or tolerance depending on timing and intensity of antigen exposure^20^. As a result, such variability may limit the extent to which maternal malaria exposure translates into consistent differences in infant vaccine responses at the population level.

In contrast, the associations of poor immunogenicity with birth outcomes (i.e., low birthweight and preterm birth) were consistent across analytical approaches. Infants born preterm or with low birthweight experienced smaller increases in serotype-specific IgG concentrations between 8 and 24 weeks and were less likely to reach the seroprotective threshold of ≥0.35 µg/mL at 24 weeks. These patterns were observed both in pooled analyses across all PCV-10 serotypes and in serotype-specific models, indicating a broad reduction in antibody acquisition rather than deficits confined to particular serotypes.

The link between adverse birth outcomes and diminished vaccine responses is biologically plausible and well supported in the literature^37^. Preterm infants exhibit relative immunologic immaturity, including reduced antigen-presenting cell function, limited T-cell help, impaired germinal center formation, and altered B-cell maturation, all of which may constrain optimal antibody production^38^. Although many preterm infants ultimately achieve protective immunity after completing vaccination schedules, early infancy may represent a period of relative vulnerability^39^. Our findings extend this understanding to pneumococcal conjugate vaccination in a malaria-endemic African setting and suggest that birth outcomes may exert a stronger influence on immunogenicity than peri-vaccination malaria exposure when clinical malaria burden is low. Our findings highlight potential for reduced early-life protection in preterm and low birthweight infants and underscore the need for further studies to evaluate whether alternative strategies (such as additional booster doses, modified schedules, or complementary maternal immunization approaches) could enhance protection in these high-risk groups.

Sensitivity analyses reinforce our primary findings. The associations between adverse birth outcomes and reduced PCV-10 responses persisted across alternative cohort definitions and analytic approaches. Conversely, the lack of association between malaria exposure or chemoprevention and vaccine responses was consistent, regardless of how malaria exposure or vaccination status was defined.

There were several limitations in our study. Although 195 infants were fully vaccinated, statistical power varied because some exposures were uncommon. For example, low birthweight (n=10) and preterm birth (n=8) were infrequent, limiting precision of effect estimates. These findings warrant investigation in larger birth cohorts with diverse adverse birth outcomes. However, the pooled GEE approach improved precision by incorporating repeated serotype-specific outcomes. Large, consistent effects for low birthweight and prematurity were detectable. Second, the predominance of asymptomatic parasitemia and rarity of febrile malaria may have limited opportunities to study malaria-related immune interference. Additionally, as this was an exploratory analysis, we chose to assess related host factors in univariate models, instead of multivariate models that account for potential interactions between covariates. We chose this approach as many host factors included in this analysis (e.g., malaria history, anemia, birth outcomes such as gestational age and birth weight) are highly inter-related and causally linked. Future studies should more deeply investigate causal links and interactions between these host factors, including gestational age and birth weight, and reduced immunogenicity and sero-conversion. Lastly, our findings are most applicable to high-transmission settings where malaria exposure during early infancy is common but often asymptomatic. Whether similar results would be observed in lower-transmission settings, where malaria episodes are less frequent but more often symptomatic or occur closer to vaccination, remains uncertain.

Overall, in this high-transmission setting, early-life malaria exposure and chemoprevention were not associated with PCV-10 immunogenicity. Instead, low birthweight and prematurity were linked to reduced antibody responses, highlighting the importance of maternal and neonatal health in optimizing vaccine effectiveness.

## Supporting information

Supplementary Material for PCV-10 manuscript

## Data Availability

The datasets generated and/or analyzed during the current study are available from the corresponding author upon reasonable request and with permission from the relevant ethics and regulatory bodies.

https://github.com/musinguzi-kenneth/PCV-10-analysis.

## Acknowledgements

We would like to thank the study participants and their families for taking part in this clinical trial and supporting our research. We thank David Vu and Angelle Desiree LaBeaud for helpful discussions and sharing reagents for bead conjugation.

## Funding Sources

This study received financial support from the National Institutes of Health (Fogarty International Center D43TW010526 (KM); U01AI155325 (PJ); T32AI165369 (AS)); Gates Foundation INV-060109 (AS, ST) and INV-092097 (MRK); the Walter V. And Idun Berry Fellowship (FB); and the Stanford Maternal and Child Health Research Institute, Woods Family Faculty Scholar in Pediatric Translational Medicine (PJ).

## Notes

### Competing Interest Statement

The authors have declared no competing interest.

### Author Declarations

This study was approved by the Stanford University Institutional Review Board, the Makerere University School of Biomedical Sciences- Research and Ethics Committee, and the Uganda National Council of Science and Technology.

