## Supplementary Material for PCV-10 manuscript for "Low birthweight and prematurity, but not malaria chemoprevention, are associated with reduced pneumococcal vaccine immunogenicity in Ugandan infants"

**Table S1:** **PCV-10 vaccination status among infants included in the immunogenicity substudy (n = 202).** Vaccination status was determined using vaccination cards and parental recall recorded at study visits. Categories reflect the number of documented PCV-10 doses received by each infant.

| PCV-10 vaccination status | n (%) |
| --- | --- |
| Received 1 dose | 3 (1.5) |
| Received 2 doses | 4 (2.0) |
| Received all 3 doses | 195 (96.5) |

**Table S2: Immunogenicity expressed as log₁₀ fold-change in serotype-specific pneumococcal IgG concentrations between 8 and 24 weeks of age following PCV-10 vaccination, showing IPTp, any parasitemia between 0-14 weeks, and PMC**. Coefficients are from pooled generalized estimating equation (GEE) models clustered by child, with serotype-specific estimates shown. Negative coefficients indicate lower fold-increase in IgG concentrations among the 3 variables. P-values are adjusted for multiple comparisons across serotypes using the Benjamini–Hochberg (BH) method. (IPTp = intermittent preventive treatment during pregnancy, PMC = perennial malaria chemoprevention).

| Serotype | IPTp | | Parasitemia between enrollment and 14 weeks | | PMC | |
| --- | --- | --- | --- | --- | --- | --- |
|  | Coefficient | Adjusted p-value | Coefficient | Adjusted p-value | Coefficient | Adjusted p-value |
| 1 | -0.017 | 0.983 | -0.68 | 0.976 | -0.171 | 0.771 |
| 4 | -0.05 | 0.983 | 0.006 | 0.986 | -0.168 | 0.771 |
| 5 | -0.011 | 0.983 | -0.509 | 0.897 | -0.131 | 0.778 |
| 6B | -0.329 | 0.848 | 0.568 | 0.897 | -0.023 | 0.954 |
| 7F | -0.178 | 0.848 | 0.241 | 0.962 | 0.292 | 0.744 |
| 9V | 0.003 | 0.983 | -0.496 | 0.897 | -0.325 | 0.735 |
| 14 | 0.199 | 0.848 | 0.098 | 0.976 | -0.379 | 0.735 |
| 18C | -0.165 | 0.848 | 0.116 | 0.976 | 0.169 | 0.771 |
| 19F | -0.103 | 0.983 | -0.227 | 0.962 | -0.419 | 0.735 |
| 23F | -0.297 | 0.848 | 0.221 | 0.962 | -0.435 | 0.735 |

**Table S3: Serotype-specific seroconversion following PCV-10 vaccination according to early-life exposures**. Odds ratios for seroconversion (defined as pneumococcal IgG concentration ≥ 0.35 µg/mL at 24 weeks of age) were estimated using pooled generalized estimating equation (GEE) logistic regression models clustered by child. Results are shown for IPTp, any parasitemia between 0-14 weeks, and PMC at the time of vaccination, with serotype-specific estimates presented. Odds ratios < 1 indicate reduced odds of seroconversion among exposed infants compared with the unexposed reference group. P-values were adjusted for multiple comparisons across serotypes using the Benjamini-Hochberg (BH) method. (IPTp = intermittent preventive treatment during pregnancy, PMC = perennial malaria chemoprevention).

| Serotype | IPTp | | Parasitemia between 0 and 14 weeks | | PMC | |
| --- | --- | --- | --- | --- | --- | --- |
|  | Odds ratio | Adjusted p-value | Odds ratio | Adjusted p-value | Odds ratio | Adjusted p-value |
| 1 | 1.03 | 0.91 | 1.15 | 0.809 | 1.61 | 0.638 |
| 4 | 0.562 | 0.323 | 2.59 | 0.468 | 1.48 | 0.638 |
| 5 | 0.74 | 0.469 | 1.22 | 0.809 | 1.74 | 0.638 |
| 6B | 0.711 | 0.323 | 2.66 | 0.43 | 0.775 | 0.638 |
| 7F | 0.897 | 0.776 | 3.12 | 0.468 | 2.42 | 0.638 |
| 9V | 1.25 | 0.538 | 3.38 | 0.468 | 1.4 | 0.638 |
| 14 | 0.665 | 0.323 | 1.77 | 0.578 | 1.34 | 0.638 |
| 18C | 0.638 | 0.323 | 3.17 | 0.43 | 1.39 | 0.638 |
| 19F | 0.675 | 0.323 | 1.9 | 0.578 | 1.93 | 0.638 |
| 23F | 0.735 | 0.323 | 2.78 | 0.43 | 1.05 | 0.875 |

**Table S4: Pooled generalized estimating equation (GEE) results for PCV-10 immunogenicity and seroconversion for only children with at least three doses documented on vaccination cards**. Immunogenicity was defined as the log₁₀ fold-change in serotype-specific pneumococcal IgG concentrations between 8 and 24 weeks of age, and seroconversion was defined as achieving a serotype-specific IgG concentration ≥0.35 µg/mL at 24 weeks of age. Pooled GEE models treated serotype-specific responses as repeated measures within participants and were clustered at the individual level. Coefficients represent mean differences in log₁₀ fold-change IgG concentrations, and odds ratios represent the odds of seroconversion. Statistical significance was assessed at α = 0.05. (HBS = hemoglobin S, IPTp = intermittent preventive treatment in pregnancy, PM = placental malaria, PMC = perennial malaria chemoprevention).

| Exposure | Immunogenicity coefficient | Immunogenicity p-value | Sero-conversion odds ratio | Sero-conversion p-value |
| --- | --- | --- | --- | --- |
| Sex | 0.008 | 0.976 | 1.10 | 0.782 |
| Low birth weight | -2.09 | p<0.001 | 0.097 | p<0.001 |
| HBS | 0.041 | 0.888 | 1.61 | 0.282 |
| Preterm birth | -1.39 | 0.059 | 0.268 | 0.049 |
| Gravidity | 0.154 | 0.647 | 1.10 | 0.830 |
| IPTp | -0.197 | 0.225 | 0.631 | 0.024 |
| PM | -0.176 | 0.529 | 0.915 | 0.802 |
| Parasitemia between enrollment and 14 weeks | -0.032 | 0.915 | 2.15 | 0.115 |
| Parasitemia between enrollment and 6 weeks | -0.212 | 0.623 | 0.244 | 0.396 |
| Anemia (at week 8) | 0.629 | 0.043 | 1.61 | 0.184 |
| PMC | 0.063 | 0.812 | 1.33 | 0.400 |

**Table S5: Pooled generalized estimating equation (GEE) results for PCV-10 immunogenicity and seroconversion for the 202 children who received at least one vaccination dose but did not complete the vaccination series**. Immunogenicity was defined as the log₁₀ fold-change in serotype-specific pneumococcal IgG concentrations between 8 and 24 weeks of age, and seroconversion was defined as achieving a serotype-specific IgG concentration ≥0.35 µg/mL at 24 weeks of age. Pooled GEE models treated serotype-specific responses as repeated measures within participants and were clustered at the individual level. Coefficients represent mean differences in log₁₀ fold-change IgG concentrations, and odds ratios represent the odds of seroconversion. Statistical significance was assessed at α = 0.05.

| Exposure | Immunogenicity coefficient | Immunogenicity  p-value | Sero-conversion odds ratio | Sero-conversion  p-value |
| --- | --- | --- | --- | --- |
| Sex | -0.232 | 0.924 | 1.17 | 0.588 |
| Low birth weight | -1.49 | 0.007 | 0.244 | 0.009 |
| HBS | 0.050 | 0.835 | 1.65 | 0.186 |
| Preterm birth | -1.36 | 0.017 | 0.266 | 0.020 |
| Gravidity | 0.185 | 0.519 | 0.881 | 0.720 |
| IPTp | -0.071 | 0.609 | 0.718 | 0.064 |
| PM | -0.215 | 0.378 | 0.802 | 0.467 |
| Parasitemia between enrollment and 14 weeks | -0.068 | 0.821 | 2.04 | 0.113 |
| Parasitemia between enrollment and 6 weeks | -0.226 | 0.594 | 2.58 | 0.361 |
| Anemia (at week 8) | 0.494 | 0.07 | 1.62 | 0.112 |
| PMC | -0.168 | 0.480 | 0.959 | 0.886 |

**Table S6: Pooled generalized estimating equation (GEE) results for PCV-10 immunogenicity and seroconversion including time since vaccination as covariate**. Immunogenicity was defined as the log₁₀ fold-change in serotype-specific pneumococcal IgG concentrations between 8 and 24 weeks of age, and seroconversion was defined as achieving a serotype-specific IgG concentration ≥0.35 µg/mL at 24 weeks of age. Pooled GEE models treated serotype-specific responses as repeated measures within participants and were clustered at the individual level. Coefficients represent mean differences in log₁₀ fold-change IgG concentrations, and odds ratios represent the odds of seroconversion. Statistical significance was assessed at α = 0.05. (HBS = hemoglobin S, IPTp = intermittent preventive treatment in pregnancy, PM = placental malaria, PMC = perennial malaria chemoprevention).

| Exposure | Immunogenicity coefficient | Immunogenicity p-value | Sero-conversion odds ratio | Sero-conversion p-value |
| --- | --- | --- | --- | --- |
| Sex | 0.021 | 0.939 | 1.250 | 0.524 |
| Low birth weight | -2.120 | 0.003 | 0.083 | 0.001 |
| HBS | 0.048 | 0.867 | 1.460 | 0.387 |
| Preterm birth | -1.380 | 0.062 | 0.217 | 0.021 |
| Gravidity | 0.138 | 0.686 | 1.260 | 0.567 |
| IPTp | -0.239 | 0.137 | 0.684 | 0.067 |
| PM | -0.209 | 0.454 | 0.980 | 0.954 |
| Parasitemia between enrollment and 14 weeks | -0.073 | 0.810 | 2.170 | 0.146 |
| Parasitemia between enrollment and 6 weeks | -0.212 | 0.628 | 1.650 | 0.647 |
| Anemia (at week 8) | 0.681 | 0.027 | 1.380 | 0.369 |
| PMC | 0.060 | 0.821 | 1.350 | 0.380 |

**Table S7: Immunogenicity and seroconversion for individual windows of vaccination**. Coefficients are from pooled generalized estimating equation (GEE) models clustered by child, with serotype-specific estimates shown. Negative coefficients indicate lower fold-increase in IgG concentrations among the 3 variables. P-values are adjusted for multiple comparisons across serotypes using the Benjamini-Hochberg (BH) method. (IPTp = intermittent preventive treatment during pregnancy, PMC = perennial malaria chemoprevention).

| Serotype | Parasitemia between enrollment and receipt of PCV1 | | Parasitemia between enrollment and receipt of PCV3 | | Parasitemia between receipt of PCV1 and PCV3 | |
| --- | --- | --- | --- | --- | --- | --- |
|  | Coefficient / odds ratio | P-value | Coefficient / odds ratio | P-value | Coefficient / odds ratio | P-value |
| Immunogenicity | -0.642 | 0.221 | -0.488 | 0.113 | -0.377 | 0.247 |
| Sero-conversion | 0.588 | 0.409 | 0.928 | 0.846 | 1.370 | 0.464 |

**Supplementary Figures**

**Figure S1:** Longitudinal serotype-specific pneumococcal IgG responses following PCV-10 vaccination, stratified by birthweight (top) and gestational age (bottom). Lines represent individual IgG concentrations for each PCV-10 serotype measured at 8 and 24 weeks of age. Panels highlight differences by <2500g (low birthweight) versus birthweight ≥2500g, and by <37 weeks (preterm) versus term birth ≥37 weeks. The horizontal dashed line on each box represents the WHO recommended IgG seroprotective cut-off of ≥0.35 µg/ml.

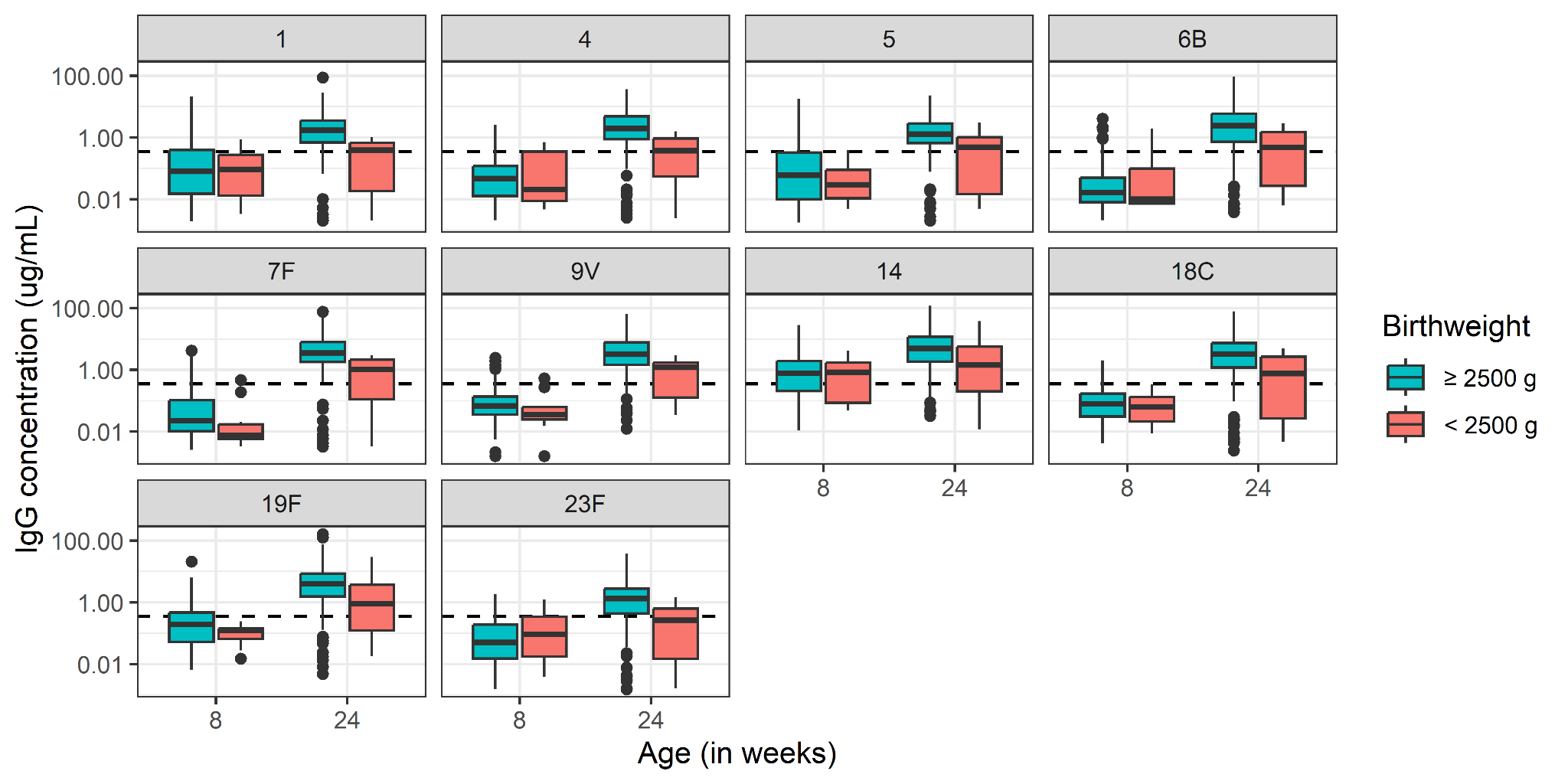

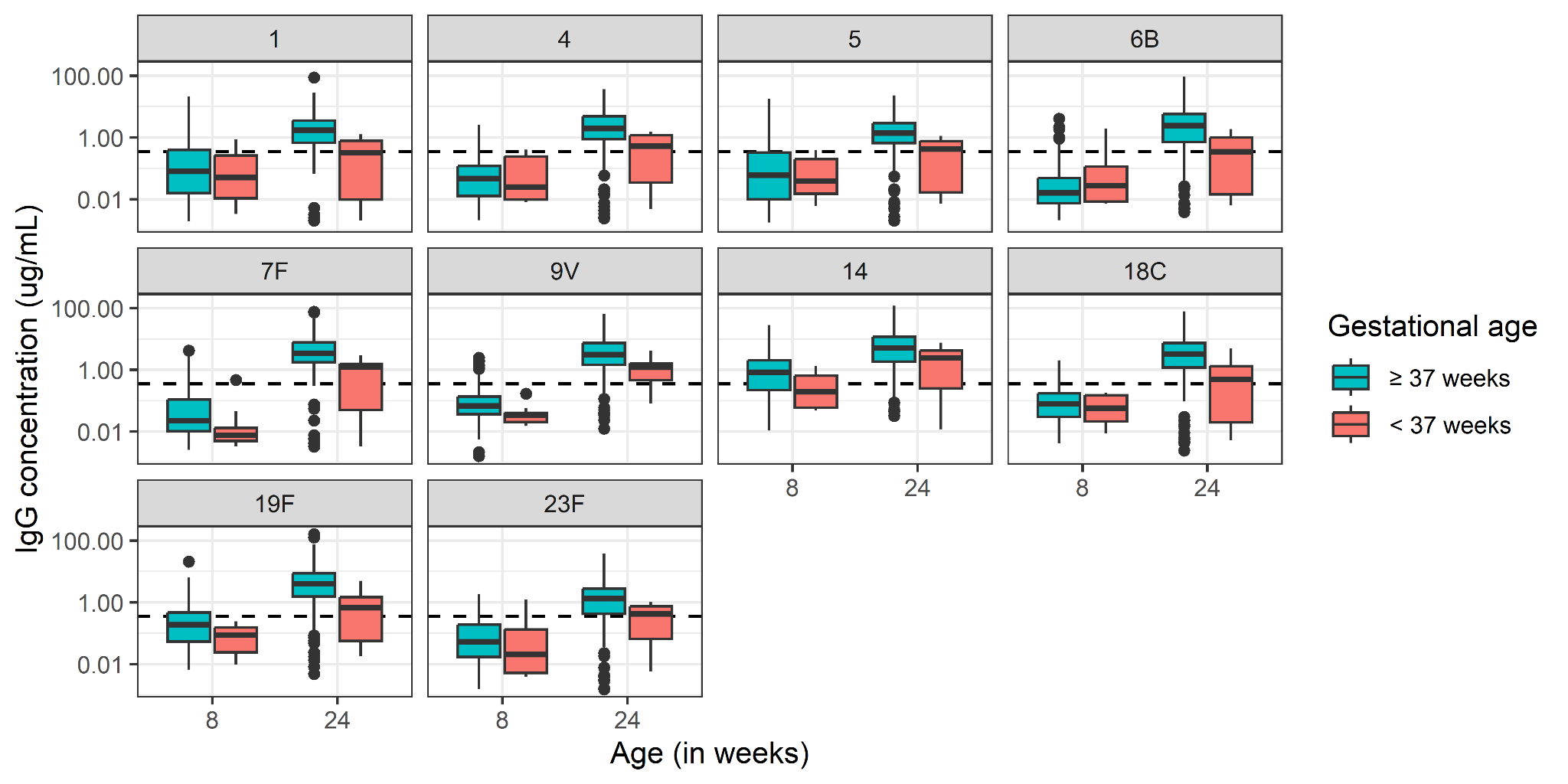

**Figure S2: Serotype-specific pneumococcal titers at weeks 8 and 24 stratified by time since receipt of PCV1 dose.** IgG concentrations (µg/mL) for each of the 10 PCV-10 serotypes are shown at 8 and 24 weeks of age, grouped according to time elapsed between receipt of the first PCV-10 dose and sample collection. Antibody concentrations were measured using a Luminex-based multiplex immunoassay and are presented on a log₁₀ scale. The horizontal dashed line represents the WHO-recommended seroprotective threshold of ≥0.35 µg/mL.

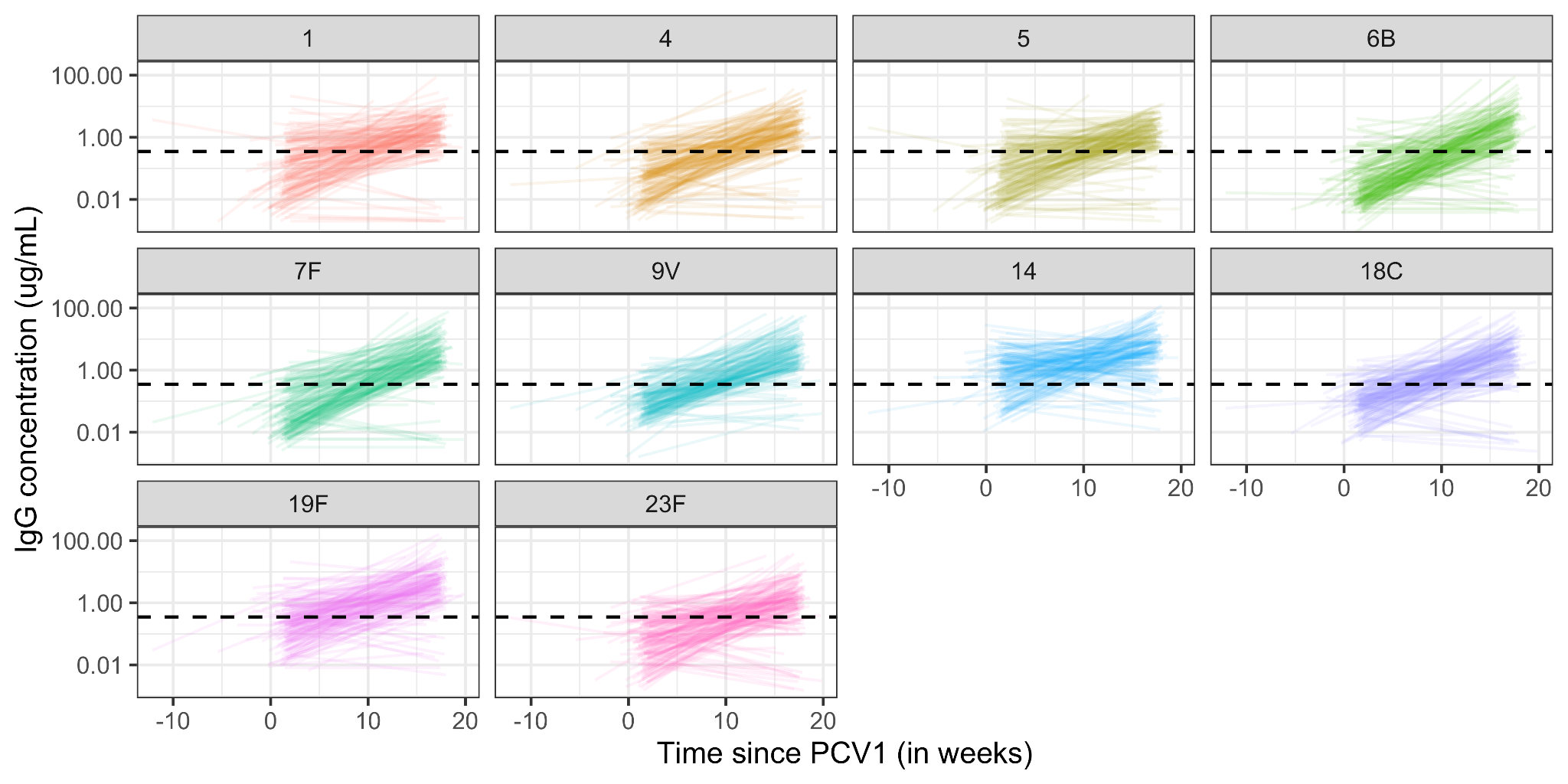
